# Risk factors for eight common cancers revealed from a phenome-wide Mendelian randomisation analysis of 378,142 cases and 485,715 controls

**DOI:** 10.1101/2023.02.15.23285952

**Authors:** Molly Went, Amit Sud, Charlie Mills, Abi Hyde, Richard Culliford, Philip Law, Jayaram Vijayakrishnan, Ines Gockel, Carlo Maj, Johannes Schumacher, Claire Palles, Martin Kaiser, Richard Houlston

## Abstract

For many cancers there are few well-established risk factors. Summary data from genome-wide association studies (GWAS) can be used in a Mendelian randomisation (MR) phenome-wide association study (PheWAS) to identify causal relationships. We performed a MR-PheWAS of breast, prostate, colorectal, lung, endometrial, oesophageal, renal, and ovarian cancers, comprising 378,142 cases and 485,715 controls. To derive a more comprehensive insight into disease aetiology we systematically mined the literature space for supporting evidence. We evaluated causal relationships for over 3,000 potential risk factors. In addition to identifying well-established risk factors (smoking, alcohol, obesity, lack of physical activity), we provide evidence for specific factors, including dietary intake, sex steroid hormones, plasma lipids and telomere length as determinants of cancer risk. We also implicate molecular factors including plasma levels of IL-18, LAG-3, IGF-1, CT-1, and PRDX1 as risk factors. Our analyses highlight the importance of risk factors that are common to many cancer types but also reveal aetiological differences. A number of the molecular factors we identify have the potential to be biomarkers. Our findings should aid public health prevention strategies to reduce cancer burden. We provide a R/Shiny app (https://mrcancer.shinyapps.io/mrcan/) to visualise findings.

## INTRODUCTION

Cancer is currently the third major cause of death with an estimated 18.1 million new cases and nearly 10 million cancer deaths in 2020^1^. By 2030 it is predicted there are likely to be 26 million new cancer cases and 17 million cancer-related deaths annually^2^. Such projections have renewed efforts to identify risk factors to inform cancer prevention programmes.

For many cancers, despite significant epidemiological research, there are few established risk factors. Although randomised-controlled trials (RCTs) are the gold standard for establishing causal relationships, they are often impractical or unfeasible because of cost, time, and ethical issues. Conversely, case-control studies can be complicated by biases such as reverse causation and confounding. Mendelian randomisation (MR) is an analytical strategy that uses germline genetic variants as instrumental variables (IVs) to infer causal relationships (**Fig. 1A**)^3^. The random assortment of these genetic variants at conception mitigates against reverse causation bias. Moreover, in the absence of pleiotropy (*i*.*e*. the presence of an association between variants and disease through additional pathways), MR can provide unconfounded disease risk estimates. Elucidating disease causality using MR is gaining popularity especially given the availability of data from large genome-wide association studies (GWAS) and well-developed analytical frameworks^3^.

**Figure 1.**
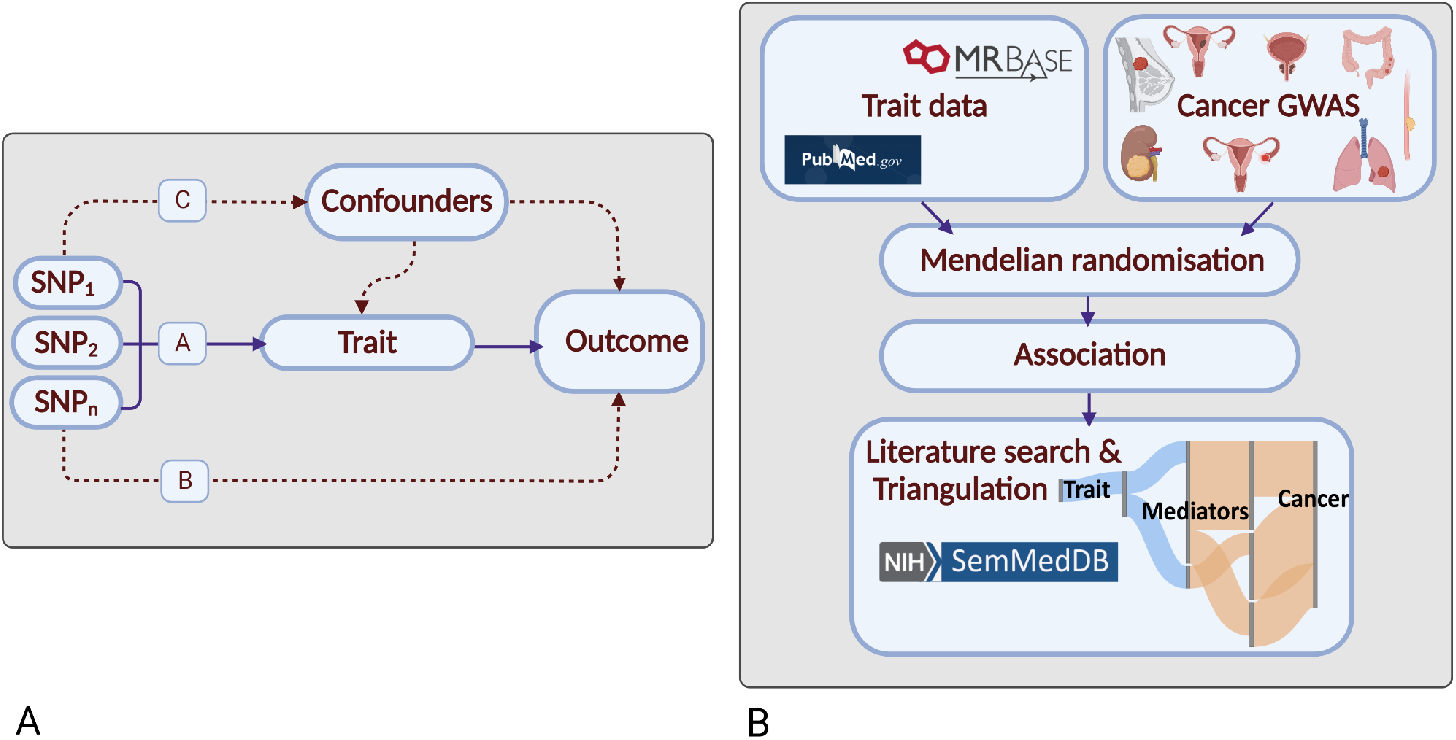
Principles of Mendelian randomisation (MR) and study overview: **(a) Assumptions in MR that need to be satisfied to derive unbiased causal effect estimates.** Dashed lines represent direct causal and potential pleiotropic effects that would violate MR assumptions. A, indicates genetic variants used as IVs are strongly associated with the trait; B, indicates genetic variants only influence cancer risk through the trait; C, indicates genetic variants are not associated with any measured or unmeasured confounders of the trait-cancer relationship. SNP, single-nucleotide polymorphism; **(b) Study overview**. Created with BioRender.com.

Most MR studies of cancer have been predicated on assumptions about disease aetiology or have sought to evaluate purported associations from conventional observational epidemiology^3,4^. A recently proposed agnostic strategy, termed MR-PheWAS, integrates the phenome-wide association study (PheWAS) with MR methodology to identify causal relationships using hitherto unconsidered traits^5^.

To identify causal relationships for eight common cancers (breast, prostate, colorectal, lung, endometrial, oesophageal, renal and ovarian), and reveal intermediates of risk, we conducted a MR-PheWAS study. We integrated findings with a systematic mining of the literature space to provide supporting evidence and derive a more comprehensive description of disease aetiology (**Fig. 1B**)^6^.

## RESULTS

### Phenotypes and genetic instruments

After filtering we analysed 3,661 traits, proxied by 336,191 genetic variants in conjunction with summary genetic data from published GWAS of breast, prostate, colorectal, lung, endometrial, oesophageal, renal, and ovarian cancers (**Table 1**; **Supplementary Table 17**). The number of SNPs used as genetic instruments for each trait ranged from one to 1,335. **Figure 2** shows the power of our MR study to identify causal relationships between each of the genetically defined traits and each cancer type. The median PVE by SNPs used as IVs for each of the 3,661 traits evaluated as risk factors was 3.4% (0.01–84%). Our power to demonstrate causal relationships *a priori* for each cancer type reflects in part inevitably the size of respective GWAS datasets (**Supplementary Table 1**).

**Table 1.**
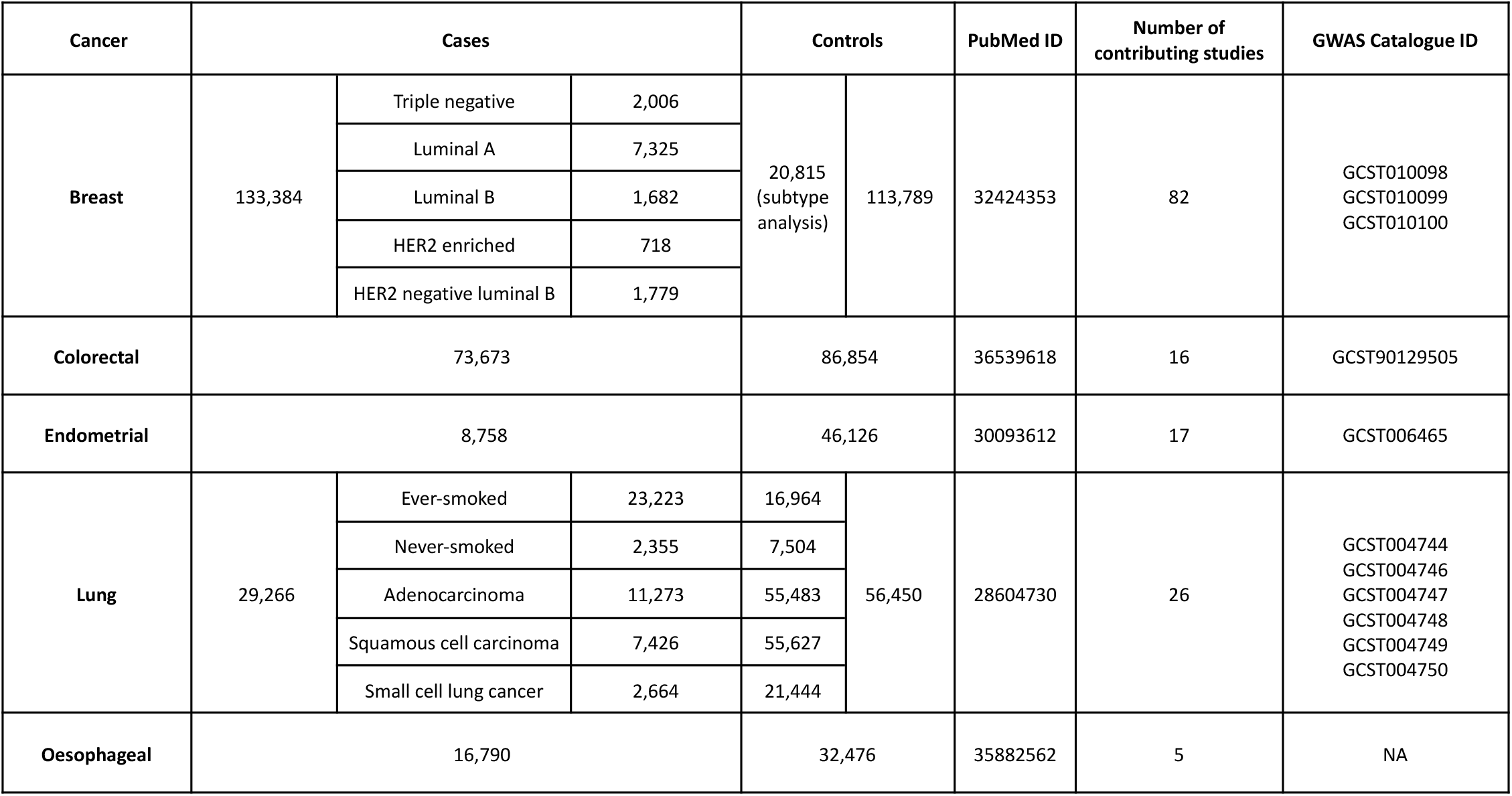
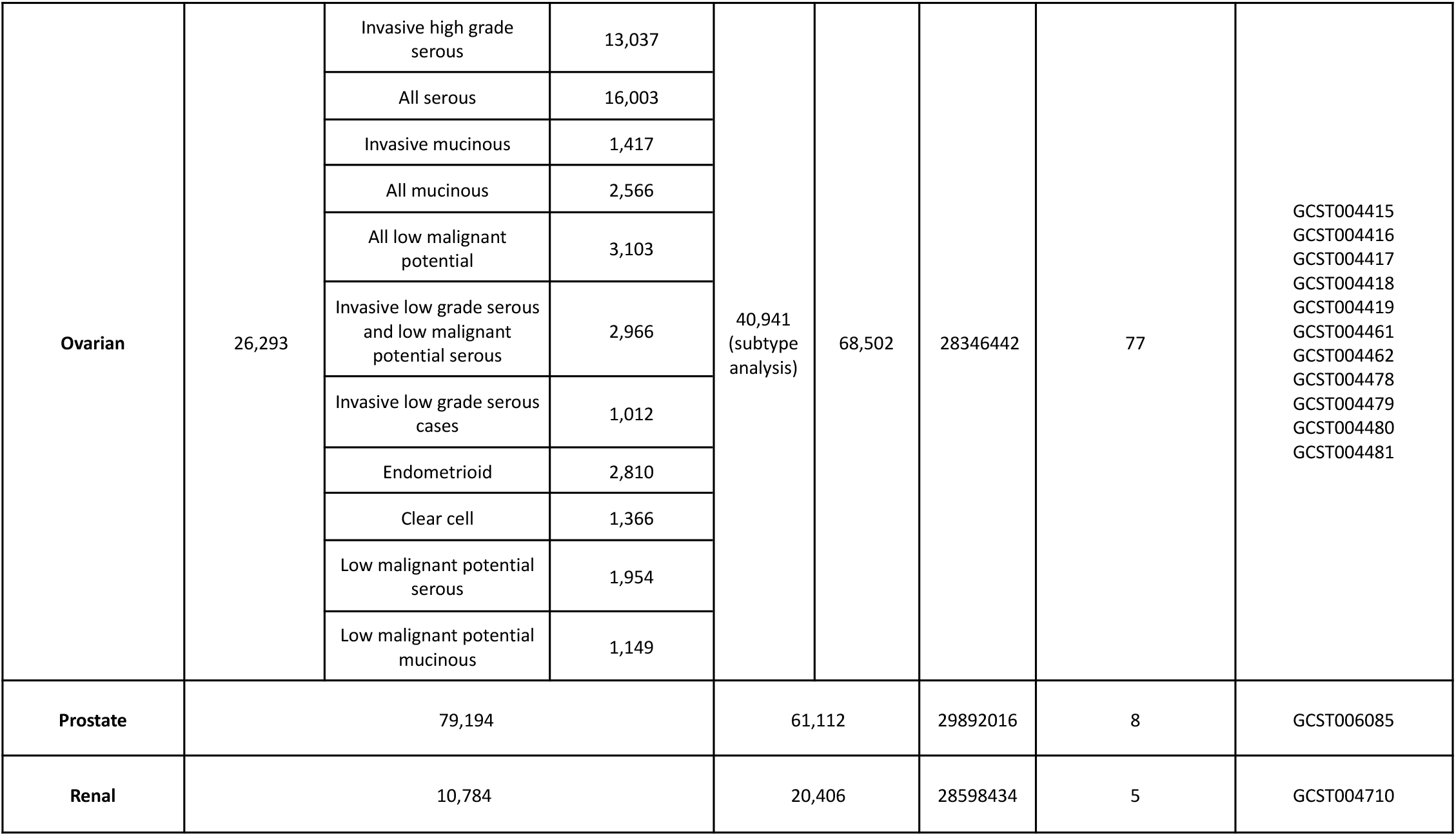
Details of cancer genome-wide association studies used in the Mendelian randomisation analysis.

**Figure 2.**
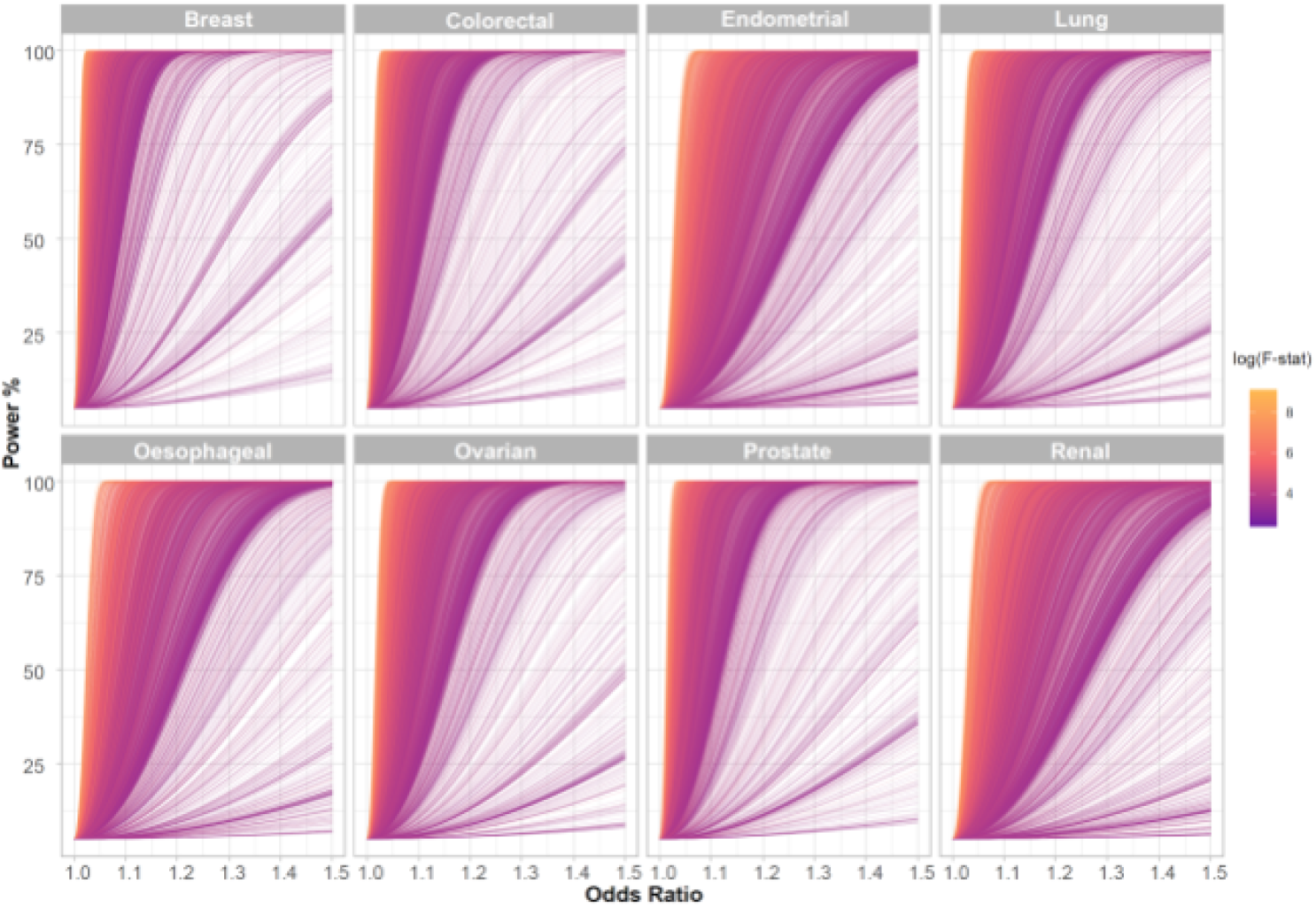
Power to demonstrate causal relationship in the Mendelian randomisation analysis across the eight different cancers. Each line represents one trait with line colour indicating F-statistic, a measure of instrument strength. The analysis of most traits is well powered across a modest range of odds ratios and this generally corresponds to those with a higher F-statistic. F-stat, F-statistic

### Causal associations identified by MR

To aid interpretation we grouped traits related to established cancer risk factors (*i*.*e*. smoking, obesity and alcohol) and those for which current evidence is inconclusive into the following categories: cardiometabolic; dietary intake; anthropometrics; immune and inflammatory factors; fatty acid (FA) and lipoprotein metabolism; lifestyle, reproduction, education and behaviour; metabolomics and proteomics; miscellaneous.

Out of the 27,066 graded associations, MR analyses provided robust evidence for a causal relationship with 123 phenotypes (0.5% of total MR analyses), 174 with probable evidence (0.6% of total), 1,652 with suggestive evidence (6% of total). Across the eight cancer types, the largest number of robust associations were observed for endometrial cancer with 37 robust associations, followed by renal cancer (n = 32), CRC (n = 21), lung (n=20), breast (n=10), oesophageal (n=3) and prostate cancer (n=1). No robust MR associations were observed for ovarian cancer (**Supplementary Table 3**).

Across all of the cancer types anthropometric traits showed the highest number of robust MR defined causal relationships (N=32; 0.1%), followed by lifestyle, reproduction, education and behaviour (N=17; 0.06%). No robust associations were observed for dietary intake or cardiometabolic categories (**Supplementary Table 3**).

To visualise the strength and direction of effect of the causal relationship between each of the traits examined and risk of each cancer type and, where appropriate, their respective subtypes we provide a R/Shiny app (https://mrcancer.shinyapps.io/mrcan/). **Fig. 3** shows a screenshot of the app for selected traits across the eight different types of cancer.

**Figure 3.**
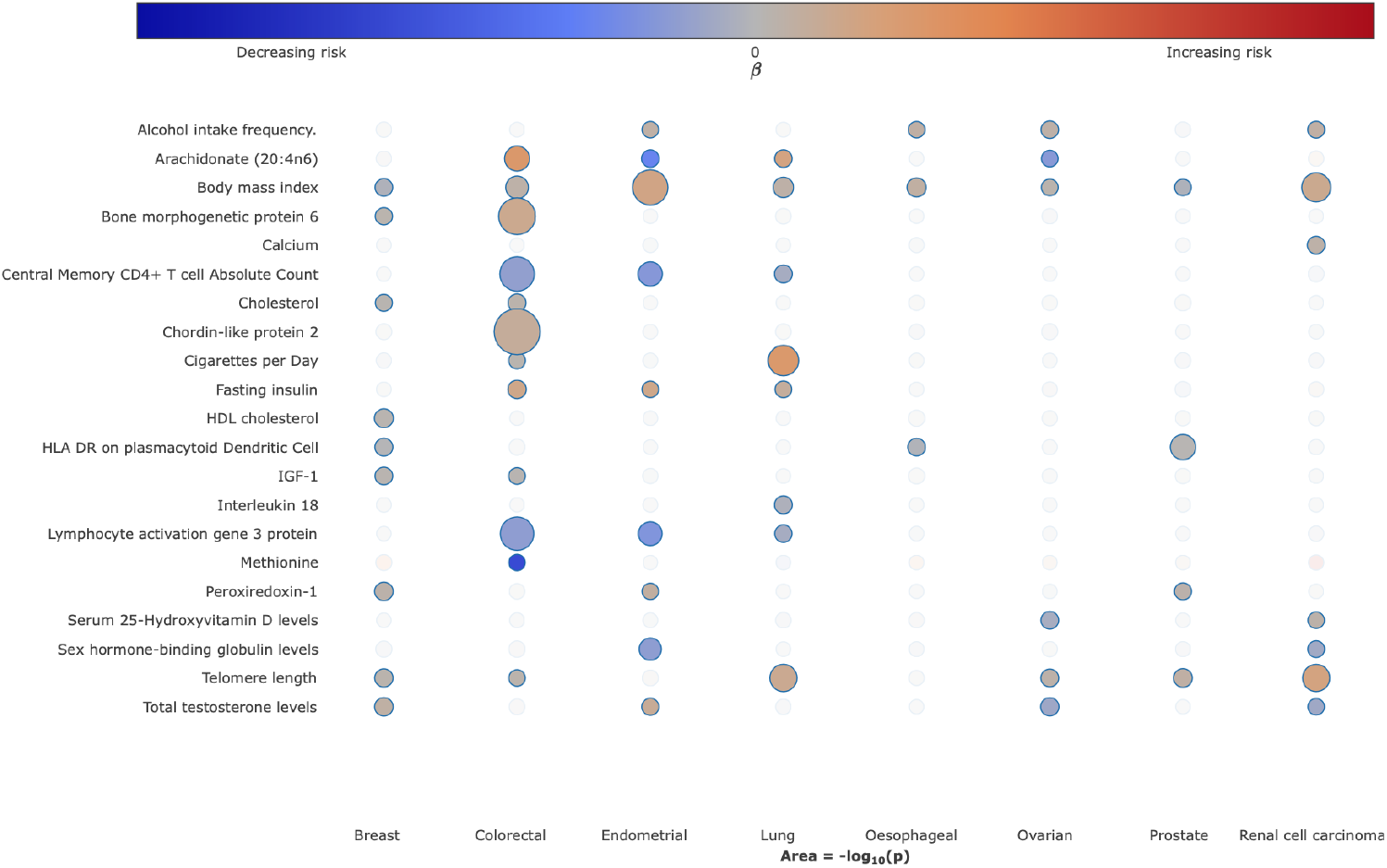
Bubble plot of the causal relationship between selected traits and risk of different cancers. Each column corresponds to cancer type. Colours on the heatmap correspond to the strength of associations (odds ratio) and their direction (red positively correlated, blue negatively correlated), the size of each node corresponding to the -log_10_ *P*-value, with increasing size indicating a smaller P-value. In the available R/Shiny app (https://mrcancer.shinyapps.io/mrcan/), moving the cursor to each bubble will reveal the underlying MR statistics.

Many of the identified causal relationships, especially those that were statistically robust or probable, have been reported in previous MR studies and are related to established risk factor categories^4,7,8^. Notably: (i) the relationship between metrics of increased body mass index (BMI) with an increased risk of colorectal, lung, renal, endometrial and ovarian cancers^9^; (ii) cigarette smoking with an increased risk of lung cancer^10^; (iii) higher alcohol consumption and increased risk of oesophageal, colorectal, lung, renal, endometrial and ovarian cancers^11^; (iv) reduced physical activity and sedentary behaviour with an increased risk of multiple cancers, including breast, lung, colorectal and endometrial^12^. As anticipated, exposure traits pertaining to cigarette smoking were not causally related to lung cancer in never smokers. Paradoxically, but as reported in previous MR analyses, increased BMI was associated with reduced risk of prostate and breast cancer, and an inverse relationship between smoking and prostate cancer risk was observed^9,13^. Our analysis also supports the reported relationship between higher levels of sex hormone-binding globulin with reduced endometrial cancer risk and a relationship between testosterone with risk of endometrial cancer and breast cancers^14,15^. Notably, exposure traits related to testosterone levels were only causally associated with luminal-A and luminal-B breast cancer subtypes.

With respect to dietary intake our analysis demonstrated probable associations between genetically predicted higher levels of coffee, oily fish, and cheese intake with reduced CRC risk and suggestive associations between genetically predicted beef and poultry intake and elevated CRC risk. We found suggestive associations between genetically predicted high serum vitamin B12 with colorectal and prostate cancer, serum calcium and 25-hydroxyvitamin-D with RCC, low blood selenium with colorectal and oesophageal cancers and methionine and zinc with reduced CRC risk. We observed no association between genetically predicted blood levels of circulating carotenoids or vitamins B6 and E for any of the cancers.

In terms of glucose homeostasis, no relationship between genetically predicted blood glucose or glycated haemoglobin was shown for any of the eight cancers. However, higher levels of genetically predicted levels of fasting insulin and insulin growth factor 1 (IGF-1) and lower proinsulin showed associations with CRC. Additionally, a suggestive association between proinsulin and renal cancer, fasting insulin and lung and endometrial cancers, and IGF-1 levels and breast cancer was observed. Amongst genetically predicted higher levels of lipoproteins, the only associations were a probable association between high density lipoprotein cholesterol (HDL-C) and breast cancer and suggestive associations between low density lipoprotein cholesterol (LDL-C) with CRC, and total cholesterol and ovarian cancer.

Genetically predicted levels of plasma FAs showed an association with reduced cancer risk. Specifically, for the omega-6 polyunsaturated FAs, lower levels of arachidonic acid (20:4n6) and gamma-linoleic acid (18:3n6) and higher levels of linoleic acid (18:2n6) and adrenic acid (22:4n6)) with reduced risk of CRC; for the omega-3 polyunsaturated FAs (alpha-linoleic acid, eicosapentaenoic acid, docosahexaenoic acid) and breast cancer risk, and arachidonic acid and endometrial cancer.

A relationship between longer lymphocyte telomere length (LTL) and an increased risk of six of the eight cancer types was identified - robust with respect to renal and lung cancers, probable for breast and prostate cancers and suggestive for colorectal and ovarian cancers.

In addition to a robust association between higher HLA-DR dendritic plasmacytoid levels and risk of prostate cancer, 26 probable associations between genetically predicted levels of other circulating immune and inflammatory factors were shown across the cancers studied. These included higher levels of IL-18 with reduced risk of lung cancer, with specificity for lung cancer in never smokers.

Our MR analysis provides support for the known relationship between colonic polyps and CRC^16^, benign breast disease and breast cancer^17^, oesophageal reflux with risk of oesophageal cancer (**Supplementary Table 13**)^18^. Other associations included possible relationships between pulmonary fibrosis and lung cancer^19^, as well as the relationship between a diagnosis of schizophrenia and lung cancer, which has been observed in conventional epidemiological studies^20^. It was noteworthy, however, that we did not find evidence to support the purported relationship between hypertension and risk of developing RCC. Similarly, our analysis did not provide evidence to support a causal relationship between either type 1 or type 2 diabetes and an increased cancer risk.

### Literature-mined support for MR causal relationships

To provide support for the associations and to gain molecular insights into the underlying biological basis of relationships we performed triangulation through systematic literature mining. We identified 55,105 literature triples across the eight different cancer types and 680,375 literature triples across the MR defined putative risk factors (**Supplementary Table 18**). Overlapping risk factor-cancer pairings from our MR analysis yielded on average 49 potential causal relationships. **Supplementary Table 19** stratifies the literature space size by trait category while recognising that causal relationships with a small literature space could be reflective of deficiencies in semantic mapping relationships with large literature spaces support triangulation. **Supplementary Table 20** provides the complete list of potential mediators for each trait. Illustrating the use of triangulation using a large literature space (defined herein as >50 triples) to support causal relationships, **Fig. 4** highlights four notable examples (IGF-1, LAG-3, IL-18, and PRDX1).

**Figure 4.**
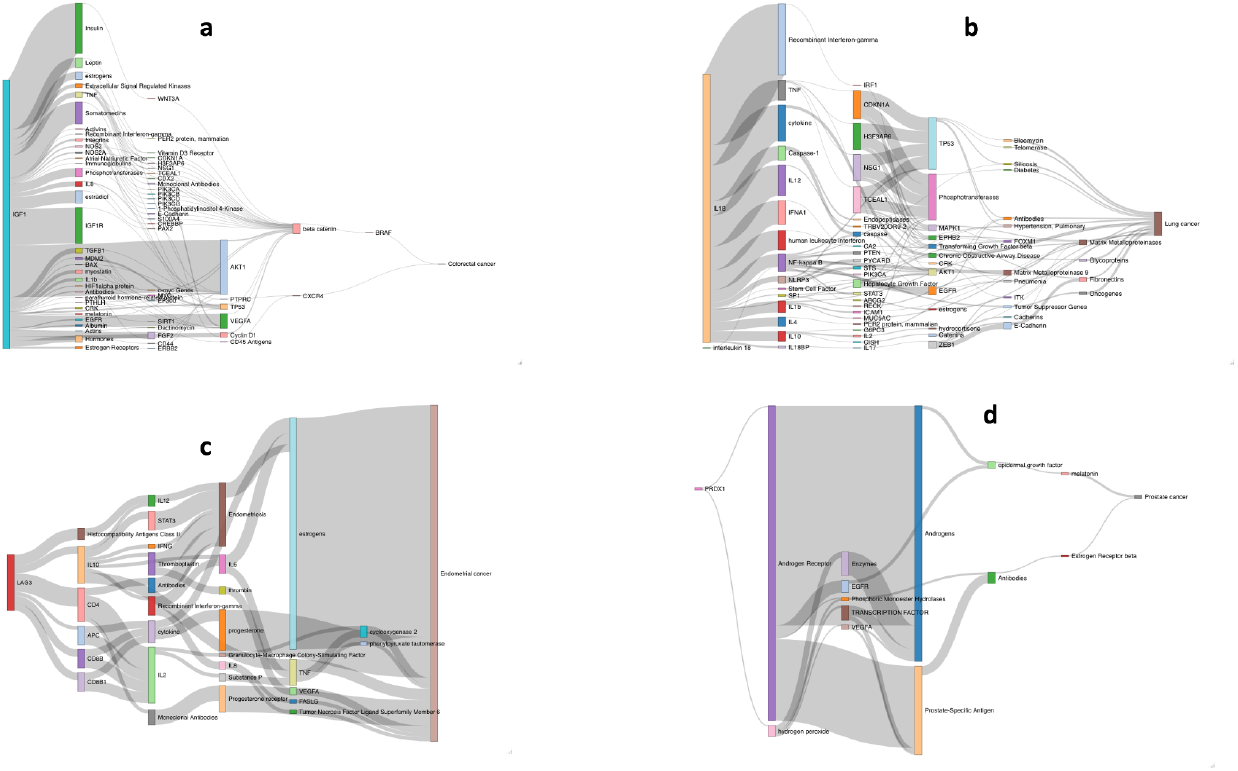
Sankey diagram of literature spaces for exemplar cancer risk factors. Relationship between: (a) *IGF-1* and colorectal cancer; (b) *IL-18* and lung cancer; (c) *LAG-3* and endometrial cancer; (d) *PRDX1* and prostate cancer.

IGF-1, which is reported to play a role in multiple cancers, appears to mediate its effect in part through beta-catenin and BRAF signalling, modulating CRC and breast cancer risk^21,22^. Whilst LAG-3 inhibition is an attractive therapeutic target in restoring T-cell function, we demonstrate genetically elevated LAG-3 levels as being associated with reduced CRC, endometrial and lung cancer. In all three of these cancers, the association appears to be at least partly mediated through IL-10 and the seemingly paradoxical relationship between LAG-3 levels and tumourgenesis suggests potentiation of T-cell function by serum LAG-3 rather than cell membrane expressed LAG-3^23^. We identify genetically predicted IL-18 levels as being associated with an increased risk of lung cancer. Our literature mining also supports a role for the decoy inhibitory protein, IL-18BP as being a mediator of lung cancer risk as well as IL-10, IL-12, IL-4 and TNF^24^. Finally, PRDX1, a member of the peroxiredoxin family of antioxidant enzymes, interacts with the androgen receptor to enhance its transactivation resulting in increased EGFR-mediated signalling and an increased prostate cancer risk^21^.

## DISCUSSION

By performing a MR-PheWAS we have been able to agnostically examine the relationship between multiple traits and the risk of eight different cancer types, restricted only by the availability of suitable genetic instruments. Importantly, many of the traits we examined have not previously been the subject of conventional epidemiological studies or been assessed by MR. Even for risk factors that were examined in many previous analyses, the number of cases and controls in our study has afforded greater power to identify potential causal associations. This has allowed us to exclude large causal effects on cancer risk for the majority of exposure traits examined in our study.

In addition to identifying causal relationships for the well-established lifestyle traits, which validates our approach, we implicate other lifestyle factors that have been putatively associated by observational epidemiology contributing to cancer risk. For example, the protective effects of physical activity, coffee, oily fish, fresh/dried fruit intake for CRC risk. A number of the causal relationships we identify have been the subject of studies of individual traits and include the association between longer LTL with increased risk of RCC and lung cancers; sex steroid hormones and risk of breast and endometrial cancer; and circulating lipids with CRC and breast cancer. Using genetic instruments for plasma proteome constituents has allowed us to identify hitherto unexplored potential risk factors for a number of the cancers, including: the cytokine like molecule, FAM3D, which plays a role in host defence against inflammation associated carcinogenesis with lung cancer^25^; the autophagy associated cytokine cardiotrophin-1 with lung, endometrial, prostate and breast cancer and the tumour progression associated antigen CD63 with endometrial cancer^26,27^. Levels of these and other plasma proteins potentially represent biomarkers worthy of future prospective studies. Clustering of MR causal effect sizes for each trait cancer relationship highlights the importance of risk factors common to many cancers but also reveal differences in their impact in part likely to be reflective of underlying biology (**Fig. 5**).

**Figure 5.**
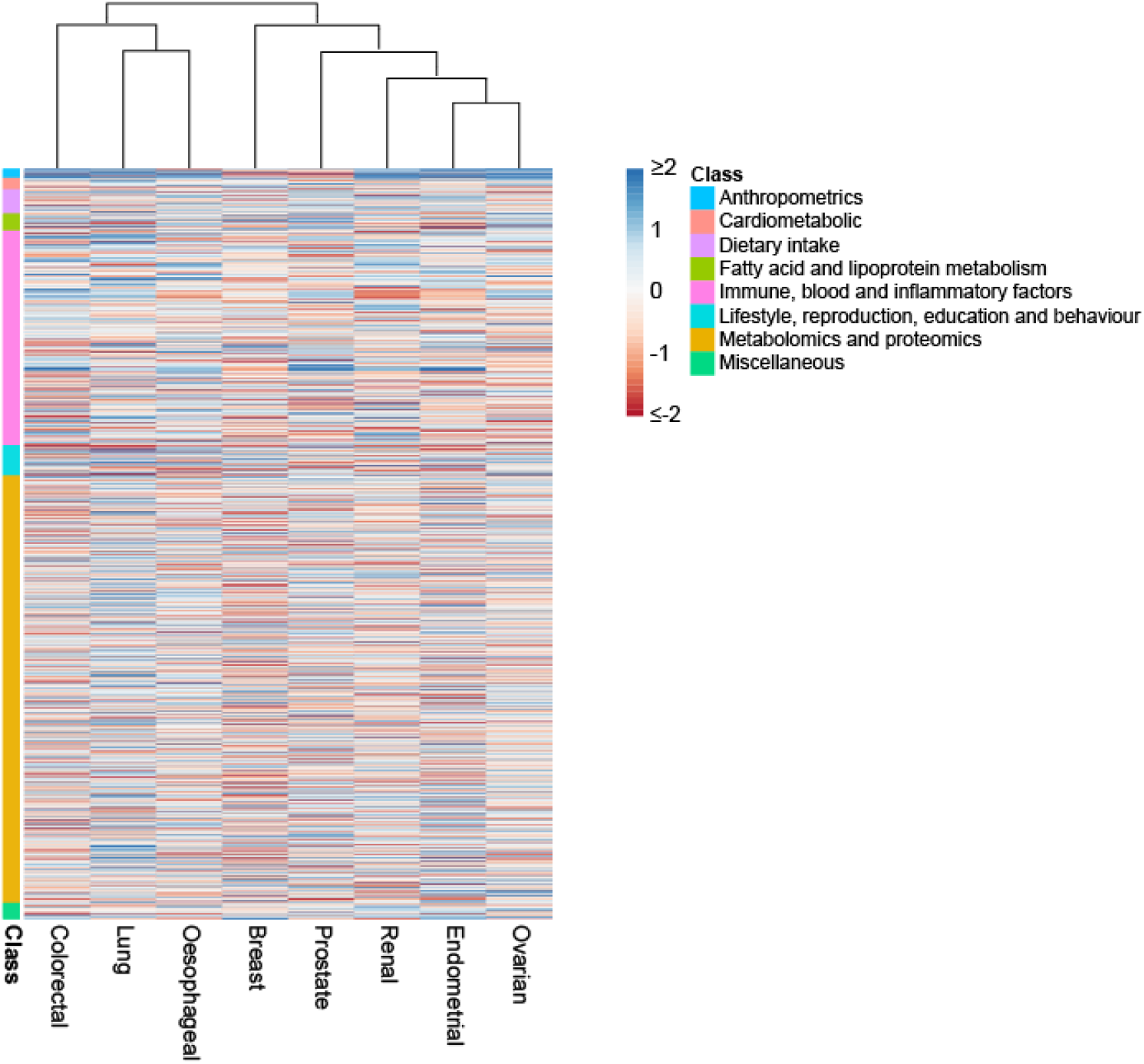
Heatmap and dendrogram showing clustering of causal associations between traits and cancer risk. Heatmap based on Z-statistics using the clustering method implemented in the pheatmap function within R. Colours correspond to the strength of associations and their direction (red positive association with risk, blue inverse association with risk). Trait classes are annotated on the left. Only traits showing an association for at least one cancer type are shown.

A principal assumption in MR is that variants used as IVs are associated with the exposure trait under investigation. We therefore used SNPs associated with exposure traits at genome-wide significance. Furthermore, only IVs from European populations were used to limit bias from population stratification. Our MR analysis does, however, have limitations. Firstly, we were limited to studying phenotypes with genetic instruments available, moreover traits such as food intake or television watching can be highly correlated with other exposures making deconvolution of the causal risk factor problematic^28,29^. Secondly, correcting for multiple testing guards against false positives especially when based on a single exposure outcome. However, the potential for false negatives is not unsubstantial. Since we have not adjusted for between trait correlations, our associations are inevitably conservative. Thirdly, for a number of traits, we had limited power to demonstrate causal associations of small effect. Fourthly, not unique to our MR analysis, is the inability of our study to deconvolute time-varying effects of genetic variants as evidenced by the relationship between obesity and breast cancer risk^30^. Finally, as with all MR studies, excluding pleiotropic IVs is challenging. To address this, we incorporated information from weighted median and mode-based estimate methods, to classify the strength of causal associations. However, there are inevitably limitations to such modelling as exemplified by the strong relationship between plasma FA and risk of CRC which has been shown to be driven by the pleiotropic *FADS* locus which has a profound effect on the metabolism of multiple FA through its gene expression^31^.

A major concern articulated regarding any MR-PheWAS is the need to provide supporting evidence from alternative sources. Herein we have sought to address this by conducting a systematic interrogation of the literature space and potentially identify intermediates to explain relationships. Although literature mined data is inevitably noisy and driven by publication bias, we have been able to provide a narrative of the causal relationships for a number of risk factors, which are attractive candidates for molecular validation.

Complementary studies are required to delineate the exact biological mechanisms underpinning associations. Our analysis does however highlight important targets for primary prevention of cancer in the population. The limited power to robustly characterise relationships between exposure traits and cancer in this study, provides an impetus for larger MR studies. Finally, we recognise that MR is not infallible and replication and triangulation of findings using different data sources, and if possible, benchmarking against RCTs is highly desirable. Such efforts could identify additional factors as targets to reduce the overall burden of cancer.

## METHODS

### Ethics approval

The analysis was undertaken using published GWAS data, hence ethical approval was not required.

### Study design

Our study had four elements. Firstly, the identification of genetic variants serving as instruments for exposure traits under investigation; secondly, the acquisition of GWAS data for the eight cancers; thirdly, MR analysis; fourthly, triangulation through literature mining to provide supporting evidence for causal relationships (**Fig. 1B**).

### Genetic variants serving as instruments

Single nucleotide polymorphisms (SNPs), considered genetic instruments, were identified from published studies or MR-Base (**Supplementary Table 1**). For each SNP, the corresponding effect estimate on a trait expressed in *per* standard deviation (SD) units (assuming a *per* allele effect) and standard error (SE) was obtained. Only SNPs with a minor allele frequency >0.01 and a trait association of *P*-values <5 × 10^−8^ in a European population GWAS were considered as instruments. We excluded correlated SNPs at a linkage disequilibrium threshold of *r*^2^ >0.01, retaining SNPs with the strongest effect. For binary traits we restricted our analyses to traits with a medical diagnosis, excluding cancer. We removed duplicate exposure traits based on manual curation.

### Cancer GWAS summary statistics

To examine the association of each genetic instrument with cancer risk, we used summary GWAS effect estimates from: (1) Online consortia resources, for breast (BCAC; https://bcac.ccge.medschl.cam.ac.uk/, accessed July 2022) and prostate cancer (PRACTICAL; http://practical.icr.ac.uk/; accessed July 2022)^32,33^; (2) GWAS catalogue (https://www.ebi.ac.uk/gwas/), for ovarian, endometrial and lung cancers (accessed September 2022)^34–36^; (3) Investigators of published work, for colorectal cancer (CRC), renal cell carcinoma (RCC) and oesophageal cancer^37–39^. Cancer subtype summary statistics were available for lung, breast and ovarian cancers. As the UK Biobank was used to obtain genetic instruments for many traits investigated, the CRC and oesophageal GWAS association statistics were recalculated from primary data excluding UK Biobank samples to avoid sample overlap bias (**Table 1**). Single nucleotide polymorphisms were harmonised to ensure that the effect estimates of SNPs on exposure traits and cancer risk referenced the same allele (**Supplementary Table 2)**^40^.

### Statistical analysis

For each SNP, causal effects were estimated for cancer as an odds ratio (OR) per SD unit increase in the putative risk factor (ORSD), with 95% confidence intervals (CIs), using the Wald ratio^41^. For traits with multiple SNPs as IVs, causal effects were estimated under an inverse variance weighted random-effects (IVW-RE) model as the primary measurement as it is robust in the presence of pleiotropic effects, provided any heterogeneity is balanced at mean zero (**Supplementary Table 3-6**)^42^. Weighted median estimate (WME) and mode-based estimates (MBE) were obtained to assess the robustness of findings (**Supplementary Table 7)**^43,44^. Directional pleiotropy was assessed using MR-Egger regression (**Supplementary Table 8**)^45^. The MR Steiger test was used to infer the direction of causal effect for continuous exposure traits (**Supplementary Table 9**)^46^. For this we estimated the proportion of variance explained (PVE) using Cancer Research UK lifetime risk estimates for each tumour type (**Supplementary Table 10**)^47^. A leave-one-out strategy under the IVW-RE model was employed to assess the potential impact of outlying and pleiotropic SNPs (**Supplementary Table 11**)^48^. Because two-sample MR of a binary risk factor and a binary outcome can be biased, we primarily considered whether there exists a significant non-zero effect, and only report ORs for consistency^49^. Statistical analyses were performed using the TwoSampleMR package v0.5.6 (https://github.com/MRCIEU/TwoSampleMR) in R (v3.4.0)^40^.

### Estimation of study power

The power of MR to demonstrate a causal relationship depends on the PVE by the instrument^50^. We excluded instruments with a F-statistic <10 since these are considered indicative of evidence for weak instrument bias^51^. We estimated study power, stipulating a *P*-value of 0.05 for each target *a priori* across a range of effect sizes as *per* Brion *et al*. (**Supplementary Table 1**)^52^. Since power estimates for binary exposure traits and binary outcomes in a two-sample setting are unreliable, we did not estimate study power for binary traits^49^.

### Assignment of statistical significance

The support for a causal relationship with non-binary traits was categorised into four hierarchical levels of statistical significance *a priori*: robust (*P*_IVW-RE_ <1.4×10^−5^; corresponding to a *P*-value of 0.05 after Bonferroni correction for multiple testing (0.05/3,500), *P*_WME_ or *P*_MBE_ <0.05, true causal direction and >1 IVs), probable (*P*_IVW-RE_ <0.05, *P*_WME_ or *P*_MBE_ <0.05, true causal direction and >1 IVs), suggestive (*P*_IVW-RE_ <0.05 or *P*_WALD_ <0.05), and non-significant (*P*_IVW-RE_ ≥0.05 or *P*_WALD_ ≥0.05) (**Supplementary Table 12)**. While non-significant associations can be due to low statistical power, they also indicate that a moderate causal effect is unlikely. For binary traits we classified associations as being supported (*P* <0.05) or not supported (*P* >0.05; **Supplementary Tables 13-16**).

### Support for causality

To strengthen evidence for causal relationships identified from the MR analysis we exploited the semantic predications in Semantic MEDLINE Database (SemMedDB), which is based on all PubMed citations^53^. Within SemMedDB pairs of terms connected by a predicate which are collectively known as ‘literature triples’ (*i*.*e*. ‘subject term 1’ – predicates – ‘object term 2’). These literature triples represent semantic relationships between biological entities derived from published literature. To interrogate SemMedDB we queried MELODI Presto and EpiGraphDB to facilitate data mining of epidemiological relationships for molecular and lifestyle traits^54–56^. For each putative risk factor-cancer pair the set of triples were overlapped and common terms identified to reveal causal pathways and inform aetiology. Based on the information profile of all literature mined triples, we considered literature spaces with >50 literature triples as being viable, corresponding to 90% of the information content^57^. We complemented this systematic text mining by referencing reports from the World Cancer Research Fund/American Institute for Cancer Research, and the International Agency for Cancer Research Global Cancer Observatory, as well as querying specific putative relationships in PubMed^58,59^.

## Data Availability

Genetic instruments can be obtained through MR-Base or from published work (Supplementary Table 1). Summary GWAS cancer data are available from: https://bcac.ccge.medschl.cam.ac.uk/bcacdata/ (breast cancer); http://practical.icr.ac.uk/blog/?page_id=8088 (prostate cancer); GWAS Catalogue ID: GCST004481 (ovarian cancer); GWAS Catalogue ID: GCST006464 (endometrial cancer); GWAS Catalogue ID: GCST004748 (lung cancer); direct communication with consortia (renal and esophageal cancers); - phs001415.v1.p1, phs001315.v1.p1, phs001078.v1.p1, phs001903.v1.p1, phs001856.v1.p1 and phs001045.v1.p1 (US based studies) and GWAS Catalog ID: GCST90129505 (European based studies) colorectal cancer.

## ACKNOWLEDGMENTS

R.S.H. acknowledges grant support from Cancer Research UK (C1298/A8362), the Wellcome Trust (214388) and Myeloma UK. A.S. is in receipt of a National Institute for Health Research (NIHR) Academic Clinical Lectureship, funding from the Royal Marsden Biomedical Research Centre, a Starter Grant from the Academy of Medical Sciences and is the recipient of the Whitney-Wood Scholarship from the Royal College of Physicians. M.K. is supported by a fellowship from the David Forbes-Nixon Foundation. We acknowledge pump-priming funding from the Royal Marsden Biomedical Research Centre Early Diagnosis, Detection and Stratified Prevention Theme. This is a summary of independent research supported by the NIHR Biomedical Research Centre at the Royal Marsden NHS Foundation Trust and the Institute of Cancer Research. The views expressed are those of the author(s) and not necessarily those of the NHS, the NIHR or the Department of Health. Support from the DJ Fielding Medical Research Trust is also acknowledged. A.H. was in receipt of a summer studentship from the Genetics Society. We thank Alex Cornish for providing code and critically appraising the manuscript.

The breast cancer genome-wide association analyses for BCAC and CIMBA were supported by Cancer Research UK (PPRPGM-Nov20\100002, C1287/A10118, C1287/A16563, C1287/A10710, C12292/A20861, C12292/A11174, C1281/A12014, C5047/A8384, C5047/A15007, C5047/A10692, C8197/A16565) and the Gray Foundation, The National Institutes of Health (CA128978, X01HG007492-the DRIVE consortium), the PERSPECTIVE project supported by the Government of Canada through Genome Canada and the Canadian Institutes of Health Research (grant GPH-129344) and the Ministère de l’Économie, Science et Innovation du Québec through Genome Québec and the PSRSIIRI-701 grant, the Quebec Breast Cancer Foundation, the European Community’s Seventh Framework Programme under grant agreement n° 223175 (HEALTH-F2-2009-223175) (COGS), the European Union’s Horizon 2020 Research and Innovation Programme (634935 and 633784), the Post-Cancer GWAS initiative (U19 CA148537, CA148065 and CA148112 - the GAME-ON initiative), the Department of Defence (W81XWH-10-1-0341), the Canadian Institutes of Health Research (CIHR) for the CIHR Team in Familial Risks of Breast Cancer (CRN-87521), the Komen Foundation for the Cure, the Breast Cancer Research Foundation and the Ovarian Cancer Research Fund. All studies and funders are listed in Zhang H et al (Nat Genet, 2020). The colorectal cancer genome-wide association analysis was supported by Ulrike Peters (GECCO), Stephanie Schmit (CCFR), Stephen Gruber (CORECT), Ian Tomlinson (CORGI, SCOT), and Malcolm Dunlop (SOCCS). Full study details and funders are listed in Fernandez-Rozadilla C et al (Nat Genet, 2023). The Prostate cancer genome-wide association analyses are supported by the Canadian Institutes of Health Research, European Commission’s Seventh Framework Programme grant agreement n° 223175 (HEALTH-F2-2009-223175), Cancer Research UK Grants C5047/A7357, C1287/A10118, C1287/A16563, C5047/A3354, C5047/A10692, C16913/A6135, and The National Institute of Health (NIH) Cancer Post-Cancer GWAS initiative grant: No. 1 U19 CA 148537-01 (the GAME-ON initiative). We would also like to thank the following for funding support: The Institute of Cancer Research and The Everyman Campaign, The Prostate Cancer Research Foundation, Prostate Research Campaign UK (now PCUK), The Orchid Cancer Appeal, Rosetrees Trust, The National Cancer Research Network UK, The National Cancer Research Institute (NCRI) UK. We are grateful for support of NIHR funding to the NIHR Biomedical Research Centre at The Institute of Cancer Research and The Royal Marsden NHS Foundation Trust. The Prostate Cancer Program of Cancer Council Victoria also acknowledge grant support from The National Health and Medical Research Council, Australia (126402, 209057, 251533,, 396414, 450104, 504700, 504702, 504715, 623204, 940394, 614296,), VicHealth, Cancer Council Victoria, The Prostate Cancer Foundation of Australia, The Whitten Foundation, PricewaterhouseCoopers, and Tattersall’s. EAO, DMK, and EMK acknowledge the Intramural Program of the National Human Genome Research Institute for their support. Genotyping of the OncoArray was funded by the US National Institutes of Health (NIH) [U19 CA 148537 for ELucidating Loci Involved in Prostate cancer SuscEptibility (ELLIPSE) project and X01HG007492 to the Center for Inherited Disease Research (CIDR) under contract number HHSN268201200008I] and by Cancer Research UK grant A8197/A16565. Additional analytic support was provided by NIH NCI U01 CA188392 (PI: Schumacher). Funding for the iCOGS infrastructure came from: the European Community’s Seventh Framework Programme under grant agreement n° 223175 (HEALTH-F2-2009-223175) (COGS), Cancer Research UK (C1287/A10118, C1287/A 10710, C12292/A11174, C1281/A12014, C5047/A8384, C5047/A15007, C5047/A10692, C8197/A16565), the National Institutes of Health (CA128978) and Post-Cancer GWAS initiative (1U19 CA148537, 1U19 CA148065 and 1U19 CA148112 – the GAME-ON initiative), the Department of Defence (W81XWH-10-1-0341), the Canadian Institutes of Health Research (CIHR) for the CIHR Team in Familial Risks of Breast Cancer, Komen Foundation for the Cure, the Breast Cancer Research Foundation, and the Ovarian Cancer Research Fund. The BPC3 was supported by the U.S. National Institutes of Health, National Cancer Institute (cooperative agreements U01-CA98233 to D.J.H., U01-CA98710 to S.M.G., U01-CA98216 toE.R., and U01-CA98758 to B.E.H., and Intramural Research Program of NIH/National Cancer Institute, Division of Cancer Epidemiology and Genetics). CAPS GWAS study was supported by the Swedish Cancer Foundation (grant no 09-0677, 11-484, 12-823), the Cancer Risk Prediction Center (CRisP; www.crispcenter.org), a Linneus Centre (Contract ID 70867902) financed by the Swedish Research Council, Swedish Research Council (grant no K2010-70X-20430-04-3, 2014-2269). PEGASUS was supported by the Intramural Research Program, Division of Cancer Epidemiology and Genetics, National Cancer Institute, National Institutes of Health.

## AUTHORSHIP

Contribution: M.W, A.S, C.M. and R.S.H designed the study. M.W, A.S., C.M., A.H., R.C., and P.L. performed statistical analyses; M.W, A.S., C.M., and R.S.H. drafted the manuscript; all authors reviewed, read, and approved the final manuscript.

## AVAILABILITY OF DATA AND MATERIAL

Genetic instruments can be obtained through MR-Base or from published work (**Supplementary Table 1**). Summary GWAS cancer data are available from: https://bcac.ccge.medschl.cam.ac.uk/bcacdata/ (breast cancer); http://practical.icr.ac.uk/blog/?page_id=8088 (prostate cancer); GWAS Catalogue ID: GCST004481 (ovarian cancer); GWAS Catalogue ID: GCST006464 (endometrial cancer); GWAS Catalogue ID: GCST004748 (lung cancer); direct communication with consortia (renal and esophageal cancers); - phs001415.v1.p1, phs001315.v1.p1, phs001078.v1.p1, phs001903.v1.p1, phs001856.v1.p1 and phs001045.v1.p1 (US based studies) and GWAS Catalog ID: GCST90129505 (European based studies) colorectal cancer.

## SUPPLEMENTARY TABLES LEGENDS

**Supplementary Table 1. List of traits examined in the Mendelian randomisation analysis and estimate of power for each trait and cancer type**.

**Supplementary Table 2. Effect allele, frequency, effect on trait and strength of association with each cancer type for SNPs used as instrumental variables**.

**Supplementary Table 3. Causal estimates from the Mendelian randomisation analysis for continuous traits and cancer risk**.

**Supplementary Table 4. Causal estimates from the Mendelian randomisation analysis for continuous traits and breast cancer subtype**.

**Supplementary Table 5. Causal estimates from the Mendelian randomisation analysis for continuous traits and lung cancer subtype**.

**Supplementary Table 6. Causal estimates from the Mendelian randomisation analysis for continuous traits and ovarian cancer subtype**.

**Supplementary Table 7. Weighted median estimate and mode-based estimates for each trait and cancer type**.

**Supplementary Table 8. MR-Egger regression analysis for each trait and cancer type. Supplementary Table 9. MR Steiger analysis for each continuous trait and cancer type**.

**Supplementary Table 10. Lifetime risk of each cancer used to calculate the proportion of variance explained**.

**Supplementary Table 11. Leave one out inverse variance weighted random-effects MR analysis for each exposure trait and cancer type**.

**Supplementary Table 12. The hierarchical levels of statistical support used to classify associations**.

**Supplementary Table 13. Causal estimates for each Mendelian randomisation method for each binary trait and cancer risk**.

**Supplementary Table 14. Causal estimates for each Mendelian randomisation method for each binary trait and breast cancer subtype**.

**Supplementary Table 15. Causal estimates for each Mendelian randomisation method for each binary trait and lung cancer subtype**.

**Supplementary Table 16. Causal estimates for each Mendelian randomisation method for each binary trait and ovarian cancer subtype**.

**Supplementary Table 17. Details of filtering applied to instrumental variables used in the Mendelian randomisation analysis**.

**Supplementary Table 18. Literature triples identified across eight different cancer types and Mendelian randomisation defined risk factors using SemMedDB**.

**Supplementary Table 19. Stratification of literature space size by trait category**.

**Supplementary Table 20. List of potential mediators for each trait identified using SemMedDB**.

